# Hypoxic Burden Predicts Symptomatic but Not Sleep Architecture Response to CPAP Therapy in Obstructive Sleep Apnea

**DOI:** 10.64898/2026.11.05.26367341

**Authors:** Sara Bäcker, Franz Ehrlich, Tony Sehr, Moritz D. Brandt, Martin Sedlmayr, Miriam Goldammer

## Abstract

The apnea-hypopnea index (AHI) remains the standard metric for assessing obstructive sleep apnea (OSA) severity, although it does not fully capture the physiological consequences of respiratory events. Hypoxic burden (HB), which integrates the frequency, depth, and duration of oxygen desaturations, has been proposed as a more comprehensive measure of OSA-related hypoxic stress. This study compared the prognostic value of HB and AHI for subjective and objective treatment outcomes in patients undergoing continuous positive airway pressure (CPAP) therapy.

Data from 412 patients with OSA who underwent diagnostic and CPAP-treated polysomnography were analyzed, including 52 patients with available follow-up questionnaires. HB was calculated using a validated event-based algorithm. Multivariable regression models were used to predict patient-reported outcomes and changes in sleep architecture, adjusted for age, sex and body mass index.

HB consistently demonstrated stronger associations with symptomatic outcomes than AHI. Models including HB showed improved model fit and explained variance. HB remained independently associated with lower follow-up fatigue and better sleep quality after adjustment for AHI. In contrast, AHI was the dominant predictor of objective sleep architecture improvements, including increases in N3 sleep, reductions in N1 sleep, and reductions in arousal index. HB provided additional independent information only for changes in arousal burden.

These findings suggest that HB and AHI capture complementary dimensions of OSA severity. While AHI primarily reflects sleep fragmentation and physiological response to CPAP, HB appears more closely related to subjective treatment benefit. Incorporating HB alongside AHI may improve clinical phenotyping and prediction of treatment response in OSA.

## 1 Introduction

Obstructive sleep apnea (OSA) is one of the most common sleep-related breathing disorders and represents a major public health burden worldwide [1]. It is characterized by recurrent episodes of partial or complete upper airway obstruction during sleep, resulting in intermittent hypoxemia, sleep fragmentation and excessive daytime sleepiness. Current estimates suggest that nearly one billion individuals worldwide are affected by OSA, with prevalence increasing due to rising obesity rates and an aging population [2]. Established risk factors include older age, obesity, male sex, craniofacial anatomy, enlarged tonsils, genetic predisposition, and lifestyle-related factors such as alcohol use and smoking [3], [4].

OSA is associated with impaired quality of life, neurocognitive dysfunction, depressive symptoms, and increased cardiovascular and metabolic risk [5]. Continuous positive airway pressure (CPAP) therapy remains the first-line treatment for moderate to severe OSA and is effective in reducing respiratory events and improving symptoms [6]. However, treatment response varies substantially between patients, and conventional measures of disease severity do not fully explain this heterogeneity [7], [8].

Traditionally, OSA severity is classified using the apnea-hypopnea index (AHI), defined as the number of respiratory events per hour of sleep [9]. Although widely used in clinical practice and guideline recommendations, the AHI has important limitations [10]. It quantifies event frequency but does not account for event duration, depth of oxygen desaturation, cumulative hypoxic stress, or the physiological consequences of respiratory disturbances [10]. As a result, patients with similar AHI values may differ considerably in symptom burden, cardiovascular risk, and therapeutic benefit [11]. Increasing criticism has therefore emerged regarding the use of AHI as the sole marker of OSA severity [12].

Hypoxic burden (HB) has been proposed by Azarbarzin et al. [13] as a more physiologically meaningful metric that quantifies the cumulative area under oxygen desaturation curves associated with respiratory events [10]. By integrating frequency, depth, and duration of desaturations, HB may better reflect the biological burden of OSA than AHI alone. Large cohort studies have shown that HB predicts cardiovascular morbidity, mortality, and adverse outcomes more strongly than traditional metrics such as AHI [14], [15]. These findings have generated growing interest in HB as a tool for risk stratification and treatment monitoring [12].

Despite this promising evidence, current research on HB has focused predominantly on cardiovascular outcomes and survival analyses [15]. Whether HB also provides clinically meaningful information for treatment decision-making, symptom improvement, and physiological response to CPAP therapy remains insufficiently studied [12].

The present study aims to systematically compare AHI and HB as predictors of both subjective and objective treatment outcomes in patients with OSA undergoing CPAP therapy. Specifically, we examined whether baseline HB provides incremental prognostic value beyond AHI for (1) patient-reported outcomes (PROs) and (2) sleep architecture. Therefore, this study seeks to clarify the potential clinical value of HB for individualized therapy decisions and treatment evaluation in OSA.

## 2 Methods

### 2.1 Dataset

This study is based on data from our local clinical sleep dataset. It was recorded in the sleep laboratory of the Department for Neurology at the University Hospital Carl Gustav Carus in Dresden. The dataset comprises routine clinical sleep studies from patients, who were referred for the evaluation of sleep disorders, including sleep-related breathing disorders, insomnia, hypersomnia, and movement disorders.

All recordings are standard in-laboratory overnight PSGs. Recordings were carried out using Alice 5 diagnostic sleep system (Philips Respironics) between 2007 and 2021 and Domino software (SOMNOmedics AG) between 2021 and 2025. The PSG data includes multimodal physiological signals, such as electroencephalography (EEG), electrooculography, electromyography, respiratory effort, airflow, oxygen saturation (SpO_2_), and electrocardiography, enabling comprehensive assessment of sleep architecture and respiratory events. CPAP therapy recordings were typically conducted one night after the diagnosis night, using a CPAP therapy device.

Sleep stages and respiratory events were scored by trained and experienced PSG technologists and somnologists according to standard clinical criteria from the AASM [16]. Diagnoses of sleep disorders were based on the International Classification of Sleep Disorders (ICSD) [17]. PROs were assessed using validated questionnaires, including the Epworth Sleepiness Scale (ESS), Fatigue Severity Scale (FSS), Pittsburgh Sleep Quality Index (PSQI), and Patient Health Questionnaire (PHQ-9). Baseline assessments were obtained at the time of diagnostic PSG night, and follow-up assessments were collected approximately one year later.

PSG recordings from different acquisition systems and file formats were imported and processed using the open-source Sleep Harmonizer framework, which is part of SleePyPhases [18]. The Sleep Harmonizer provides a unified interface for loading and harmonizing polysomnographic recordings and annotations across multiple vendor-specific formats, enabling standardized downstream analyses.

For the present analysis, we included all patients diagnosed with obstructive sleep apnea who underwent diagnostic night and CPAP therapy nights within three days of each other, both of which were monitored using PSG. Only cases with available covariate data were included, resulting in a total of 412 cases. For the analysis of symptomatic outcomes, we additionally only included patients with good CPAP adherence and with available follow-up questionnaire data. Good CPAP adherence was defined as an average daily use of the CPAP device of at least 4 hours and treatment effectiveness. This resulted in a total of 52 cases with PROs (see Figure 1).

**Figure 1:**
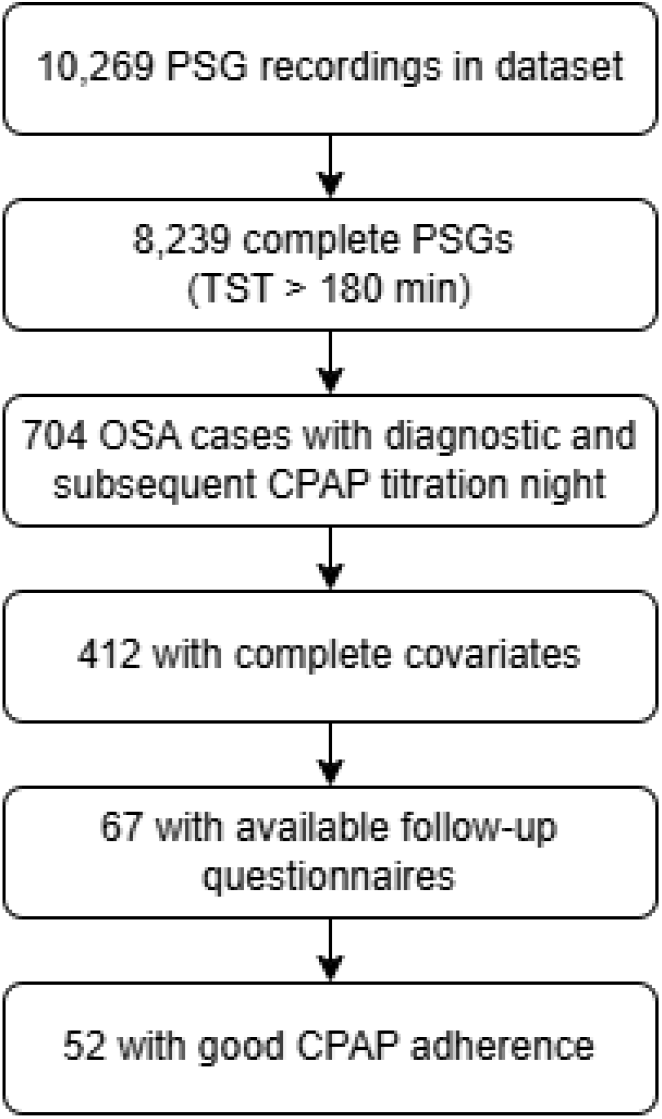
Flowchart of cohort selection of the used clinical sleep dataset.

Demographic and clinical covariates included in all regression models are age, sex, and body mass index (BMI), due to their established association with obstructive sleep apnea severity and clinical outcomes. The characteristics of the dataset used can be found in Table 1.

**Table 1:** Demographic characteristics, patient-reported outcomes, and polysomnographic outcomes of the investigated patients with obstructive sleep apnea. Values are presented as mean ± standard deviation unless otherwise specified.

| **Demographic characteristics (n = 412)** |  | **Value** |
|---|---|---|
| Age (years) |  | 59.37 ± 13.94 |
| Age range (years) |  | 21 - 92 |
| Male sex (%) |  | 66.75 |
| BMI (kg/m<sup>2</sup>) |  | 30.28 ± 5.62 |
| **Patient-reported outcomes (n = 52)** | **Baseline** | **Follow-up** |
| ESS score | 8.90 ± 4.93 | 7.98 ± 4.75 |
| FSS score | 32.53 ± 16.96 | 29.35 ± 15.64 |
| PSQI score | 6.67 ± 3.36 | 5.91 ± 3.01 |
| PHQ-9 score | 6.67 ± 4.51 | 6.12 ± 4.41 |
| **Polysomnographic outcomes (n = 412)** | **Diagnostic night** | **Therapy night** |
| N3 (%) | 13.12 ± 9.42 | 22.23 ± 10.17 |
| N1 (%) | 31.95 ± 19.82 | 18.51 ± 12.43 |
| REM latency (min) | 158.59 ± 91.62 | 103.53 ± 75.22 |
| Arousal index (events/h) | 30.84 ± 20.11 | 18.41 ± 12.65 |

Abbreviations: BMI, body mass index; ESS, Epworth Sleepiness Scale; FSS, Fatigue Severity Scale; PSQI, Pittsburgh Sleep Quality Index; PHQ-9, Patient Health Questionnaire.

### 2.2 Calculation of Sleep Apnea Severity Metrics

The hypoxic burden (HB), as defined by Azarbarzin et al. [13], describes the cumulative impact of oxygen desaturations associated with respiratory events. It is calculated as the sum of the areas between a pre-event baseline and the oxygen saturation curve, associated with each apnea and hypopnea event. The HB is designed to integrate both the depth and duration of each desaturation event, thereby providing a more comprehensive picture of hypoxic stress than simple event counts such as AHI.

The algorithm for HB calculation was implemented as follows. Individual SpO_2_ segments associated with manually scored respiratory events are averaged and processed to calculate the search window. The boundaries of the search window are determined by the two peaks before and after the end of the event of the averaged SpO_2_ curve. The pre-event baseline for each respiratory event is defined as the maximum SpO_2_ value within 120 seconds prior to the end of the event. The area between the baseline and the SpO_2_ curve is calculated over the subject-specific search window for each event. The HB is the cumulative sum of the individual areas, divided by the total sleep time.

We implemented the HB algorithm in Python, based on the MATLAB implementation by Philip de Chazal [19]. We adopted it for different sampling rates and optimized the filter function for the averaged SpO_2_ signal by using a lowpass FIR filter with cutoff frequency of 0.1 Hz to smooth the signal. Our implementation can be found under the MIT License on GitLab, see https://gitlab.com/sleep-is-all-you-need/hypoxic-burden.

We evaluated and tested our HB implementation thoroughly on the Sleep Heart Health Study (SHHS) dataset [21], comprising nearly 5,800 polysomnographic recordings. The dataset was provided by the National Sleep Research Resource (NSRR) [22]. The implemented HB algorithm showed excellent agreement with the reference implementation in MATLAB by Philip de Chazal. This was demonstrated by near-perfect Pearson correlation (r > 0.999) and minimal systematic bias (bias = −1.15 %min/h).

However, the definition and selection of the respiratory events used as input for the algorithm have a substantial impact on the resulting HB values. We observed that the annotated apnea and hypopnea events in the SHHS polysomnography data do not directly correspond to the events used to calculate the specified AHI value in the SHHS dataset files. Although there are quite a few different AHI definitions, the number of annotated respiratory events exceeds the values included in all of these definitions. This is most likely because all scored apnea and hypopneas events are listed in the annotations, and even basic criteria for AHI calculation are applied afterwards. For instance, if an apnea was scored during a wake epoch, it would still be listed in the respiratory event list but not considered in any AHI calculation. While this is conclusive in itself, it creates the additional task of event selection for event-based algorithms.

Therefore, a harmonized event selection was required to ensure consistency between calculation of HB and AHI. We replicated the AHI according to the definition “ahi_a0h3” from the SHHS dataset. The AHI was defined as the total number of apneas and hypopneas divided by the total sleep time in hours. For apneas, the criteria were a ≥ 90 % drop in airflow for ≥ 10 seconds without oxygen desaturation threshold and with or without arousals. For hypopneas, the criteria were a ≥ 30 % drop in airflow for ≥ 10 seconds, as well as a ≥ 3 % oxygen desaturation from a pre-event baseline with or without arousals. Oxygen desaturation was computed relative to a pre-event baseline, defined as the maximum of a 120-second window preceding event onset. Additionally, the calculation only includes apneas and hypopneas that began during sleep.

After automatically applying these filtering criteria to the annotated respiratory events, the resulting AHI values showed strong agreement with the SHHS reference values. The comparison between the implemented AHI and the *ahi_a0h3* reference demonstrated a very high correlation (Pearson r = 0.981, p < 0.001), with a small systematic bias of −0.50 events/h, indicating a slight underestimation relative to the reference method, most likely due to slight implementation differences of the event selection rules.

Using the same respiratory event definitions, HB and AHI were calculated for our local clinical dataset.

### 2.3 Statistical Analysis

Multivariable linear regression analyses were performed to evaluate the incremental prognostic value of HB beyond the AHI in predicting symptomatic and physiological responses to CPAP treatment in patients with OSA.

The two outcome domains, PROs and PSG-derived outcomes, were analyzed. PROs included daytime sleepiness (ESS), fatigue (FSS), sleep quality (PSQI), and depressive symptoms (PHQ-9). For these analyses, follow-up questionnaire scores obtained approximately one year after diagnostic night served as dependent variables while adjusting for baseline questionnaire scores. PSG-derived outcomes included N3 sleep (%), N1 sleep (%), REM latency, and arousal index. For PSG-derived outcomes, treatment effects were quantified as the difference between the CPAP therapy night and the diagnostic night:

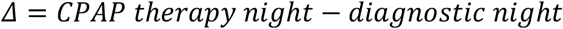

Separate hierarchical regression models were calculated for each outcome. For PSG-derived outcomes, Model A included diagnostic-night AHI and the predefined covariates age, sex, and BMI. Model B included diagnostic-night HB and the same covariates. Model AB included both AHI and HB in addition to the covariates.

The model structure for PSG-derived outcome parameters is:

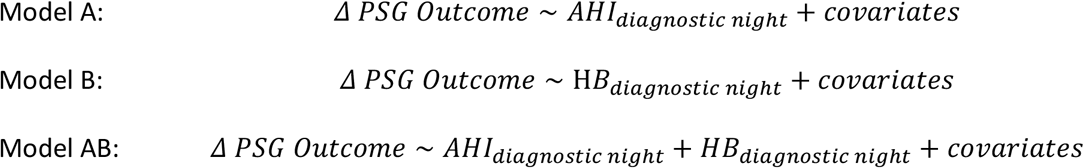

For PRO analyses, follow-up questionnaire scores were modeled as the dependent variable while adjusting for the corresponding baseline questionnaire score (*PRO*_*follow*−*up*_ ~ *PRO*_*baseline*_). This analysis of covariance approach was chosen instead of analyzing change scores (*Δ PRO) be*cause it accounts for baseline differences between participants, reduces the influence of regression to the mean, and generally provides greater statistical efficiency. The resulting model structures were:

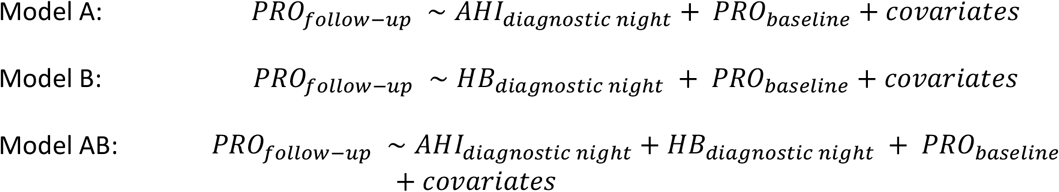

Accordingly, regression coefficients for the PRO models represent the association between baseline OSA severity and follow-up PRO scores after adjustment for the corresponding baseline questionnaire score and covariates.

Model performance was evaluated using the coefficient of determination (R^2^), reflecting explained variance, and the Akaike Information Criterion (AIC), which assesses model fit while penalizing model complexity. Comparisons across Models A, B, and AB were used to determine the relative predictive value of AHI and HB. The inclusion of both predictors in Model AB enabled evaluation of whether HB contributed unique explanatory information after accounting for AHI, while also allowing assessment of the independent associations of each OSA severity metric with treatment outcomes.

Model assumptions were assessed using residual diagnostics, including tests for normality (Omnibus and Jarque–Bera tests). Due to deviations from normality in several models, robust standard errors (HC1) were calculated as sensitivity analyses. Results were considered robust if statistical significance remained consistent. Statistical tests were two-sided, and a p-value < 0.05 was considered statistically significant. All statistical analyses were performed using Python’s statsmodels library. Continuous variables are presented as mean ± standard deviation, unless otherwise stated.

## 3 Results

In the following, the results of the HB and AHI during diagnostic and therapy night, as well as the multivariable linear regression models for PROs and PSG-derived outcomes are depicted.

### 3.1 Hypoxic Burden and Apnea Hypopnea Index

Descriptive statistics and correlations of the derived sleep apnea severity metrics for all OSA patients in our local clinical dataset are presented in Table 2. Both AHI and HB decreased markedly from the diagnostic night to the CPAP therapy night. AHI and HB were strongly correlated in both the diagnostic night (r = 0.80, p < 0.001) and the therapy night (r = 0.82, p < 0.001).

**Table 2:** Calculated sleep apnea severity metrics apnea-hypopnea index (AHI) and hypoxic burden (HB) in the used clinical dataset (n = 412) with mean and standard deviation as well as pearson correlations (r) for diagnostic and therapy night.

| **Sleep Apnea Severity** | **Diagnostic night** | **Therapy night** |
|---|---|---|
| AHI (events/h) | 22.29 ± 22.31 | 5.50 ± 7.55 |
| HB (%min/h) | 60.00 ± 82.72 | 11.66 ± 22.09 |
| **Correlations** |  |  |
| Correlation of AHI and HB | r = 0.80 (p < 0.001) | r = 0.82 (p < 0.001) |

### 3.2 Patient-Reported Outcomes

A broadly consistent pattern emerged across all investigated PROs. Models including HB generally showed better model fit than models including AHI alone, as reflected by higher R^2^ values and lower AIC values. Results of the multivariable linear regression models for PROs are summarized in Table 3.

**Table 3:** Results of the multivariable linear regression models for the patient-reported outcomes (PROs) at follow-up, including daytime sleepiness (ESS), fatigue (FSS), sleep quality (PSQI) and depressive symptoms (PHQ-9).

| Outcome | Modell | AHI β (p) | HB β (p) | R² | AIC |
|---|---|---|---|---|---|
| **ESS follow-up<br>(Daytime sleepiness)** | A | **-0.075 (0.003)** | — | 0.694 | 256.7 |
|  | B | — | **-0.017 (0.001)** | 0.709 | **254.0** |
|  | AB | 0.019 (0.772) | -0.020 (0.125) | **0.710** | 255.9 |
| **FSS follow-up<br>(Fatigue)** | A | -0.085 (0.137) | — | 0.821 | 347.4 |
|  | B | — | **-0.026 (0.023)** | 0.832 | 343.9 |
|  | AB | 0.263 (0.080) | **-0.074 (0.015)** | **0.843** | **342.4** |
| **PSQI follow-up<br>(Sleep quality)** | A | -0.030 (0.131) | — | 0.565 | 237.5 |
|  | B | — | **-0.009 (0.023)** | 0.591 | 234.2 |
|  | AB | 0.089 (0.093) | **-0.026 (0.018)** | **0.617** | **232.9** |
| **PHQ-9 follow-up<br>(Depression)** | A | -0.031 (0.078) | — | 0.795 | 224.3 |
|  | B | — | **-0.007 (0.041)** | 0.800 | **223.1** |
|  | AB | 0.018 (0.713) | -0.010 (0.276) | **0.801** | 224.9 |

Note: Significant regression coefficients and improved model fit indices are highlighted in bold. Included covariates are age, sex, BMI and baseline PRO. Abbreviations: AHI, apnea-hypopnea index; HB, hypoxic burden; β, beta coefficients; p, p-values; R<sup>2</sup>, coefficient of determination; AIC, Akaike Information Criterion; ESS, Epworth Sleepiness Scale; FSS, Fatigue Severity Scale; PSQI, Pittsburgh Sleep Quality Index; PHQ-9, Patient Health Questionnaire.

In the AHI-only models (Model A), higher AHI at diagnostic night was associated with lower follow-up ESS scores (β = −0.075, p = 0.003), whereas no significant associations were observed for fatigue (FSS), sleep quality (PSQI), or depressive symptoms (PHQ-9). In contrast, higher HB at diagnostic night was significantly associated with lower follow-up symptom burden across all investigated PROs in the HB-only models (Model B), including ESS (β = −0.017, p = 0.001), FSS (β = −0.026, p = 0.023), PSQI (β = −0.009, p = 0.023), and PHQ-9 (β = −0.007, p = 0.041).

When both metrics were entered simultaneously (Model AB), associations between AHI and follow-up symptom burden were no longer statistically significant. In contrast, HB remained independently associated with lower follow-up fatigue (β = −0.074, p = 0.015) and better sleep quality (β = −0.026, p = 0.018), whereas associations with daytime sleepiness and depressive symptoms were attenuated and no longer significant after adjustment for AHI.

Overall, models including HB demonstrated superior explanatory performance compared with models including AHI alone. The largest improvements in model fit were observed for fatigue and sleep quality, for which the combined models achieved the highest R^2^ values and lowest AIC values for these outcomes (Table 3).

Figure 2 shows the adjusted regression coefficients and 95% confidence intervals from the fully adjusted models. HB demonstrated consistently negative effect estimates across all PROs, indicating a tendency toward lower residual symptom burden with increasing baseline HB. In contrast, AHI estimates were associated with wider confidence intervals and none remained statistically significant after adjustment for HB.

**Figure 2:**
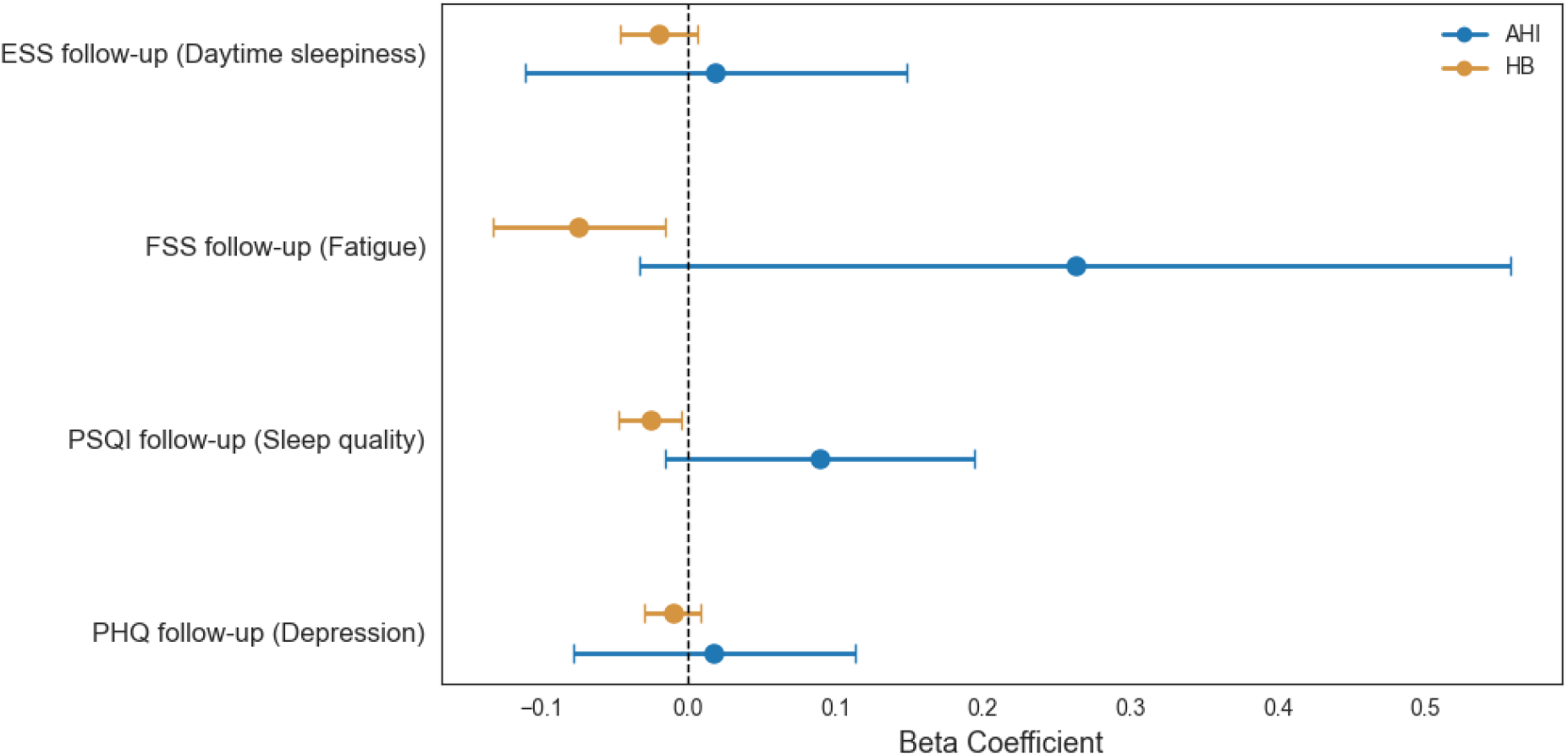
Forest plot of the adjusted regression coefficients (β) with 95% confidence intervals from the fully adjusted multivariable regression models (Model AB) for the four PRO outcomes. Shown are the independent effects of apnea-hypopnea index (AHI) and hypoxic burden (HB) at diagnostic night, each adjusted for baseline PRO, age, sex, and BMI.

### 3.3 PSG-Derived Outcomes

AHI at diagnostic night was consistently associated with CPAP-related improvements in sleep architecture, higher AHI at diagnostic night was associated with stronger improvements in sleep architecture at therapy night. Although similar associations were observed for HB, models including AHI generally demonstrated better explanatory performance than the corresponding HB-only models. Results of the multivariable linear regression models for PSG-derived outcomes are summarized in Table 4.

**Table 4:** Results of the multivariable linear regression models for PSG-derived outcomes including changes in N3 sleep, N1 sleep, REM latency, and arousal index.

| Outcome | Modell | AHI β (p) | HB β (p) | R² | AIC |
|---|---|---|---|---|---|
| **Δ N3 (%)** | A | **0.161 (<0.001)** | — | 0.125 | **3058** |
|  | B | — | **0.038 (<0.001)** | 0.098 | 3070 |
|  | AB | **0.133 (<0.001)** | 0.010 (0.321) | **0.127** | 3059 |
| **Δ N1 (%)** | A | **-0.310 (<0.001)** | — | 0.209 | **3423** |
|  | B | — | **-0.070 (<0.001)** | 0.146 | 3445 |
|  | AB | **-0.275 (<0.001)** | -0.012 (0.426) | **0.210** | 3424 |
| **Δ REM Latency** | A | **-0.933 (<0.001)** | — | 0.055 | **4961** |
|  | B | — | **-0.242 (<0.001)** | 0.052 | 4962 |
|  | AB | -0.600 (0.101) | -0.016 (0.240) | **0.058** | 4961 |
| **Δ Arousal Index** | A | **-0.400 (<0.001)** | — | 0.228 | 3480 |
|  | B | — | **-0.099 (<0.001)** | 0.195 | 3497 |
|  | AB | **-0.291 (<0.001)** | **-0.038 (0.019)** | **0.239** | **3476** |

Note: Significant regression coefficients and improved model fit indices are highlighted in bold. Included covariates are age, sex and BMI. Abbreviations: AHI, apnea-hypopnea index; HB, hypoxic burden; β, beta coefficients; p, p-values; R<sup>2</sup>, coefficient of determination; AIC, Akaike Information Criterion.

In the AHI-only models (Model A), higher baseline AHI predicted larger increases in N3 sleep (β = 0.161, p < 0.001), larger reductions in N1 sleep (β = −0.310, p < 0.001), shorter REM latency (β = −0.933, p < 0.001), and greater reductions in arousal index (β = −0.400, p < 0.001). Similar associations were observed in the HB-only models (Model B), including N3 sleep (β = 0.038, p < 0.001), N1 sleep (β = − 0.070, p < 0.001), REM latency (β = −0.242, p < 0.001), and arousal index (β = −0.099, p < 0.001), although model performance was consistently lower than in the corresponding AHI models.

When both metrics were entered simultaneously (Model AB), AHI remained independently associated with increases in N3 sleep (β = 0.133, p < 0.001), reductions in N1 sleep (β = −0.275, p < 0.001), and reductions in arousal index (β = −0.291, p < 0.001). In contrast, the corresponding HB effects were substantially attenuated and no longer statistically significant. For REM latency, neither AHI nor HB remained independently associated after mutual adjustment.

Overall, inclusion of HB provided little additional explanatory value beyond AHI for most PSG-derived outcomes. The notable exception was the arousal index, for which both AHI (β = −0.291, p < 0.001) and HB (β = −0.038, p = 0.019) retained independent associations in the combined model. Accordingly, the combined model achieved the highest explained variance (R^2^ = 0.239) and the lowest AIC (3476) for this outcome.

Figure 3 displays the adjusted regression coefficients and 95% confidence intervals from the fully adjusted models. AHI showed consistent associations with CPAP-related changes in sleep architecture, particularly for N3 sleep, N1 sleep, and arousal index. In contrast, HB coefficients were generally small and centered near zero, with confidence intervals overlapping the null effect for most outcomes. The only independent HB association was observed for the arousal index.

**Figure 3:**
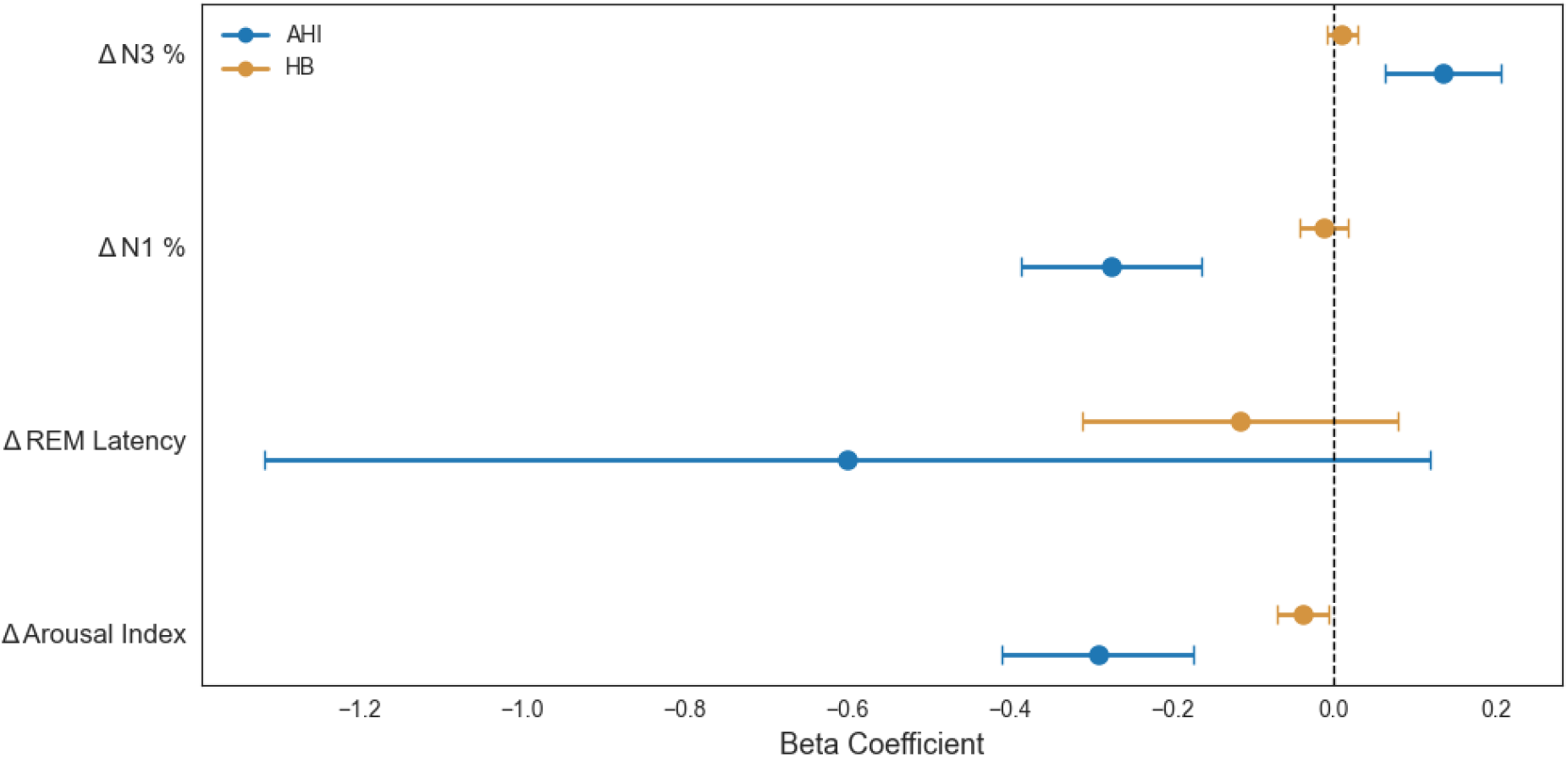
Forest plot of the adjusted regression coefficients (β) with 95% confidence intervals from the fully adjusted regression models (Model AB) for the PSG-derived outcomes. Shown are the independent effects of apnea-hypopnea index (AHI) and hypoxic burden (HB) at diagnostic night, each adjusted for age, sex, and BMI.

## 4 Discussion

In this study, we assessed the incremental value of HB for OSA treatment. Therefore, we compared the prognostic value of HB with AHI for both PROs and PSG-derived parameters. As the main finding, HB consistently provided clinically relevant information beyond AHI for symptomatic outcomes, whereas AHI remained more strongly related to changes in sleep architecture as a response to CPAP therapy. These results suggest that AHI and HB reflect complementary pathophysiological dimensions of OSA.

### 4.1 Hypoxic Burden and Symptomatic Treatment Response

Across multiple PROs, including daytime sleepiness, fatigue, sleep quality, and depressive symptoms, HB emerged as the more consistent independent predictor of follow-up symptom burden than AHI. Models including HB generally demonstrated better fit than models including AHI alone, and associations between AHI and symptomatic outcomes disappeared after adjustment for HB. This pattern indicates that the apparent relationship between AHI and symptom improvement is largely driven by the hypoxic component of respiratory events rather than event frequency itself.

Baseline questionnaire scores were the strongest predictors of follow-up values, resulting in overall high explained variance (R^2^ values 0.565-0.843). Adding HB to the models consistently improved model fit, as reflected by higher R^2^ and lower AIC values compared with models including AHI alone.

From a clinical perspective, these findings support the concept that OSA severity and expected treatment benefits should not be assessed solely by the number of respiratory events. While AHI quantifies event frequency, it does not adequately capture the physiological burden imposed by repeated oxygen desaturations. HB may therefore represent the clinically more relevant dimension of subjective disease severity, particularly with regard to daytime functioning and treatment-related symptom improvement.

This was particularly evident for fatigue and sleep quality, where higher baseline HB was associated with lower residual symptom burden at follow-up. These findings suggest that patients with greater nocturnal hypoxic stress showed lower residual symptom burden following CPAP therapy after adjustment for baseline symptom severity. This suggests that HB may identify patients more likely to experience favorable symptomatic response to CPAP.

Our findings support the clinical relevance of HB as a marker of disease severity and treatment response, as HB is a stronger predictor of symptomatic outcomes, such as daytime sleepiness and fatigue. This aligns with prior evidence from pediatric populations, where HB was shown to correlate more closely with symptom burden than the AHI [23].

### 4.2 PSG-Derived Outcomes: Complementary Information from AHI

HB was associated with sleep architecture changes in single-predictor models. However, after adjustment for AHI, most associations disappeared, indicating that these effects were largely shared with respiratory event frequency. AHI remained the dominant predictor of sleep architecture response, particularly increases in N3 sleep, reductions in N1 sleep, shorter REM latency, and lower arousal burden. In contrast, HB provided additional information only for changes in arousal burden.

The consistent pattern of best R^2^ for Model AB, but best AIC for Model A reflects, that the improvements in sleep architecture are *best* explained (R^2^) by the combination of AHI and HB, but *simplest* explained (AIC) by just AHI.

These findings suggest that respiratory event frequency primarily reflects the mechanical burden of repetitive airway obstruction and associated sleep fragmentation. Patients with higher AHI may therefore show greater immediate normalization of sleep continuity once obstructive events are eliminated during CPAP. By comparison, HB is less relevant to predict these acute changes in sleep structure.

Additionally, regression models were carried out for sleep microstructure parameters such as the spindle index and delta power. However, no statistically significant results could be obtained for either AHI or HB, and the model performance was very poor for these outcomes. Therefore, they were not reported in detail in this article.

Taken together, our results indicate that AHI and HB should not be viewed as competing metrics, but rather as complementary measures. By that interpretation, AHI characterizes fragmentation-related sleep disruption, whereas HB represents the physiological consequences of intermittent hypoxemia and their relationship to subjective clinical symptoms.

### 4.3 Clinical Implications

Current clinical classification of OSA relies predominantly on AHI thresholds. However, patients with similar AHI values often differ substantially in symptom burden, cardiovascular risk, and treatment response [11], [13]. Our findings add to the growing evidence that metrics incorporating hypoxic stress may improve phenotyping and risk stratification.

HB showed independent incremental value in multivariable models and may help identify patients more likely to benefit symptomatically from CPAP therapy. This could be particularly relevant for individuals with inconsistent clinical presentations, such as a high symptom burden despite a moderate AHI, or limited symptoms despite a severe AHI.

Accordingly, future approaches to OSA assessment may benefit from integrating both event frequency and hypoxic burden rather than relying on AHI alone.

### 4.4 Limitations

Several limitations should be considered. Firstly, the number of available follow-up questionnaires was comparatively small, resulting in a small sample size for PRO analyses (n = 52), which limits statistical power and generalizability. Therefore, these findings should be interpreted as exploratory and require replication in larger and more diverse cohorts. In contrast, analyses of PSG-derived outcomes were conducted in a substantially larger cohort (n = 412).

One limitation relates to the scoring of hypopneas. According to the AASM criteria, hypopneas may be defined either by a ≥ 3 % oxygen desaturation or by an associated arousal. In the present study, hypopneas were scored based on the oxygen desaturation criterion only, whereas arousal-related hypopneas without accompanying desaturation were not included. This may have led to an underestimation of respiratory events compared with full AASM scoring. However, identical apnea and hypopnea events were used for the calculation of both AHI and HB, ensuring internal methodological consistency across metrics.

Regarding the HB calculation, no universally accepted standard algorithm currently exists. Different computational approaches have been proposed, limiting comparability across studies and currently restricting routine clinical implementation [10], [12]. Furthermore, clinically meaningful threshold values for HB have not yet been clearly defined, which currently constrains direct translation into treatment algorithms or severity classification.

Existing HB algorithms may incompletely capture certain respiratory event characteristics, particularly prolonged events or complex desaturation patterns. Although the algorithm is designed to capture the depth and duration of desaturations, it calculates a patient-specific search window that averages the length of the desaturations and defines a uniform duration for each patient. Consequently, longer events are not fully captured.

In addition, HB depends on accurate scoring of respiratory events and therefore remains influenced by manual event annotation. In contrast, alternative approaches such as time under 90 % oxygen saturation (T90) or accumulated desaturation area metrics quantify nocturnal hypoxemia directly from the oximetry signal. An ideal metric would capture hypoxic stress independently of manually annotated respiratory events.

Finally, HB is highly correlated with AHI, which may complicate simultaneous modeling and limit interpretability in multivariable analyses due to collinearity.

## 5 Conclusion

HB improved prediction of several symptomatic outcomes as a response to CPAP treatment, particularly fatigue and sleep quality, whereas AHI remained the dominant predictor of short-term sleep architecture changes. These findings support the interpretation that respiratory event frequency and hypoxic burden capture distinct but complementary aspects of OSA severity. Incorporating HB alongside AHI may improve clinical phenotyping and better explain heterogeneity in treatment response.

## 6 Acknowledgements

The authors would like to thank the Data Integration Center of the University Hospital Carl Gustav Carus Dresden at the Faculty of Medicine of the Technical University of Dresden for providing the data utilised in this study. The Data Integration Center is funded by the Network University Medicine under the German Federal Ministry of Education and Research (grant number NUM-DIZ: 01KX2524).

The authors would like to express their gratitude to Christian Strassberger for discussion on details of the hypoxic burden algorithm, and to Michael Arzt for discussion of clinical relevance and clinical covariates.

The Sleep Heart Health Study (SHHS) was supported by National Heart, Lung, and Blood Institute cooperative agreements U01HL53916 (University of California, Davis), U01HL53931 (New York University), U01HL53934 (University of Minnesota), U01HL53937 and U01HL64360 (Johns Hopkins University), U01HL53938 (University of Arizona), U01HL53940 (University of Washington), U01HL53941 (Boston University), and U01HL63463 (Case Western Reserve University). The National Sleep Research Resource was supported by the National Heart, Lung, and Blood Institute (R24 HL114473, 75N92019R002).

## Data and Code Availability Statement

The dataset is available from the Data Integration Center of the University Hospital Carl Gustav Carus Dresden at the Faculty of Medicine of the Technical University of Dresden upon reasonable request, but subject to applicable ethical and data protection regulations. Service fees may apply.

The code for the hypoxic burden algorithm is publicly available under the MIT License on GitLab: https://gitlab.com/sleep-is-all-you-need/hypoxic-burden.

## Ethics Approval Statement

This study was approved by the ethics committee of the University of Duisburg-Essen (25-12624-BO) and acknowledged by the local ethics committee of TUD Dresden University of Technology (BO-EK-171042026), permitting the use of anonymized data without informed consent.

